# Incidence of unruptured intracranial aneurysms: A nation-wide population based longitudinal study

**DOI:** 10.64898/2026.11.12.26367153

**Authors:** Daniel Jaraj, Ahmed Ali, Per Persson, Johan Ljungqvist, Magnus Tisell

## Abstract

**Background:** Understanding the epidemiology of unruptured intracranial aneurysm (UIAs) is crucial for risk stratification, clinical decision making, and public health planning. The aim of this study was to perform a nationwide population-based analysis to examine the incidence of unruptured intracranial aneurysms in Sweden.

**Methods:** Data were obtained from the Swedish National Patient Register for all persons, aged 18 years and older, diagnosed with an intracranial aneurysm (ICD-10: I67.1) between the years 2000 and 2020. Crude incidence was calculated for the total population and age- and gender-stratified groups. Age standardized incidence was assessed using the European standard population. The incidence of intracranial aneurysms diagnosed with concurrent subarachnoid hemorrhage (SAH) was also examined.

**Results:** During the study period, 8681 patients were diagnosed with UIA. The median age of diagnosis was 63 years and 65.1% were women. The age-standardized incidence increased approximately 74% % between 2000 and its peak: from 4.6 (95% CI: 4,26–4.95) per 100,000 in the year 2000 to 7.99 (95% CI: 6.7–7.54) per 100,000 in the year 2019. □ The increase in incidence of UIAs was most pronounced among women and those aged above 60. During the study period, the number of cases with SAH decreased slightly.

**Conclusions:** The increasing incidence of UIAs in Sweden might be due improved clinical detection rates, demographic changes, and changes in the prevalence of risk factors. These findings highlight the importance of enhanced surveillance, effective risk stratification, and targeted public health strategies to address the growing burden of UIAs.

## BACKGROUND

Aneurysmal subarachnoid hemorrhage (aSAH) is the most severe form of stroke. Because of its high mortality rate and risk of major sequelae among survivors it poses a substantial burden on both individuals and society [1-3].

So far much of the epidemiological research on intracranial aneurysms has been focused on the outcome, namely subarachnoid hemorrhage (SAH) [4]. In contrast, data on unruptured intracranial aneurysms (UIAs) remain comparatively limited. Furthermore, most previous studies of UIAs have relied on cross-sectional prevalence estimates derived from smaller samples, selected populations, or screening cohorts, while large-scale longitudinal data at the population level are more scarce [4, 5].

Incidence studies on hospital-based diagnoses probably underestimate the true occurrence of disease in the population, compared to e.g. screening studies. However, the main advantage of hospital-based incidence studies is that they better reflect the diagnosed disease burden that drives clinical follow-up, imaging surveillance, and preventive interventions. Moreover, in contrast to screening studies, nationwide registry data enables the assessment of temporal trends and comparisons across regions and healthcare systems, in ways that are difficult to achieve with smaller cohort studies. While the incidence of SAH has decreased substantially over the past decades, it is less clear whether the frequency of UIAs has followed a similar trend[3, 6]. There is a lack of large-scale longitudinal data shedding light on this issue. Up-to-date population data spanning an extended period are therefore crucial for a better understanding of the current epidemiology of UIAs, as well as their implications for healthcare planning, resource allocation, and risk stratification. The aim of this study was therefore to examine the nation-wide incidence of UIAs during a 20-year period in Sweden.

## METHODS

### Study population and data sources

This nationwide population-based study was conducted using data from the Swedish National Patient Register (NPR) and Statistics Sweden (SCB), the national agency for official population statistics in Sweden.[7-9] The NPR has complete nationwide coverage of all inpatient hospital admissions since 1987 and specialised outpatient care since 2001. It contains individual-level data linked through a unique personal identity number and includes information on diagnoses coded according to the International Classification of Diseases, Tenth Revision (ICD-10), dates of admission and discharge, and healthcare provider characteristics. Annual population counts stratified by age and sex were obtained from SCB. Because the Swedish healthcare system is publicly funded with universal access, almost all hospital diagnoses of intracranial aneurysms are captured in the NPR dataset. The NPR is maintained by the Swedish National Board of Health and Welfare, a centralized government agency, and has been extensively validated demonstrating high diagnostic accuracy [7, 9, 10].

### Case definition and inclusion criteria

Using ICD-10 codes, we identified all individuals aged ≥18 years with a diagnosis of intracranial aneurysm (I67.1) recorded in the NPR between 1 January 2000 and 31 December 2020. Both inpatient and outpatient diagnoses were included.

To restrict the analyses to unruptured intracranial aneurysms (UIAs), individuals whose first recorded diagnosis of intracranial aneurysm occurred in conjunction with subarachnoid haemorrhage (SAH) were excluded from the main incidence analyses.

SAH was identified using ICD-10 code I60. Individuals with a recorded diagnosis of intracranial aneurysm preceding a subsequent SAH event were classified as having an unruptured aneurysm at baseline and were included in the UIA cohort. Individuals with an SAH diagnosis (I60) without a concurrent aneurysm diagnosis (I67.1) were not included in this study.

### Statistical analysis

Crude incidence rates of UIA were calculated as the number of incident cases per calendar year divided by the corresponding Swedish population aged ≥18 years, expressed per 100 000 person-years. Age-specific incidence rates were calculated separately for men and women in 10-year age groups.

To account for changes in the age structure of the population over time and to allow comparison with other populations, age-standardized incidence rates were calculated using the direct standardization method [11] using the European standard population as reference [12]. The European standard population categorizes individuals in 5-year age groups. Therefore, in order to age-standardized the present sample we excluded those aged < 20 years (n=25) for the analysis of the standardized incidence. However, for the age-specific incidence rates all individuals were included. 95% Confidence Intervals were calculated assuming a Poisson distribution.

Temporal trends in the incidence of UIA and SAH were analyzed using Poisson regression models, with calendar year entered as a continuous variable and the logarithm of the population size included as an offset term. Model assumptions were assessed, including evaluation of overdispersion. All analyses were made using R software for statistical computing. [13] Main R packages utilized were tidyverse, ggplot2 and epitools. [14-16]

### Ethical approval

The study was approved by the Swedish Ethical Review Authority (dnr 2021-01805) and conducted in accordance with the Helsinki declaration.

### Patient and public involvement statement

The study is a retrospective epidemiological registry-based study where data were obtained from the Swedish National Patient Register. The data provide largely complete coverage of inpatient and outpatient care in Sweden and span two decades. Given the nature of the study’s design, neither patients nor the general public were involved.

## RESULTS

Between 2000 and 2020, a total of 9,535 incident cases of intracranial aneurysms were diagnosed. After excluding individuals with concurrent SAH at diagnosis, 8,681 persons with UIAs remained. Of these, 65.1% were women (n = 5,648) and 34.9% men (n = 3,033). The median age at diagnosis was 63 years (IQR 19), with women being slightly older than men (63 vs. 62 years). The total number of women was consistently higher in all age groups particularly among the oldest. The age and gender distribution of individuals diagnosed with UIA during the study period is shown in table 1 and figure 1.

**Table 1.**
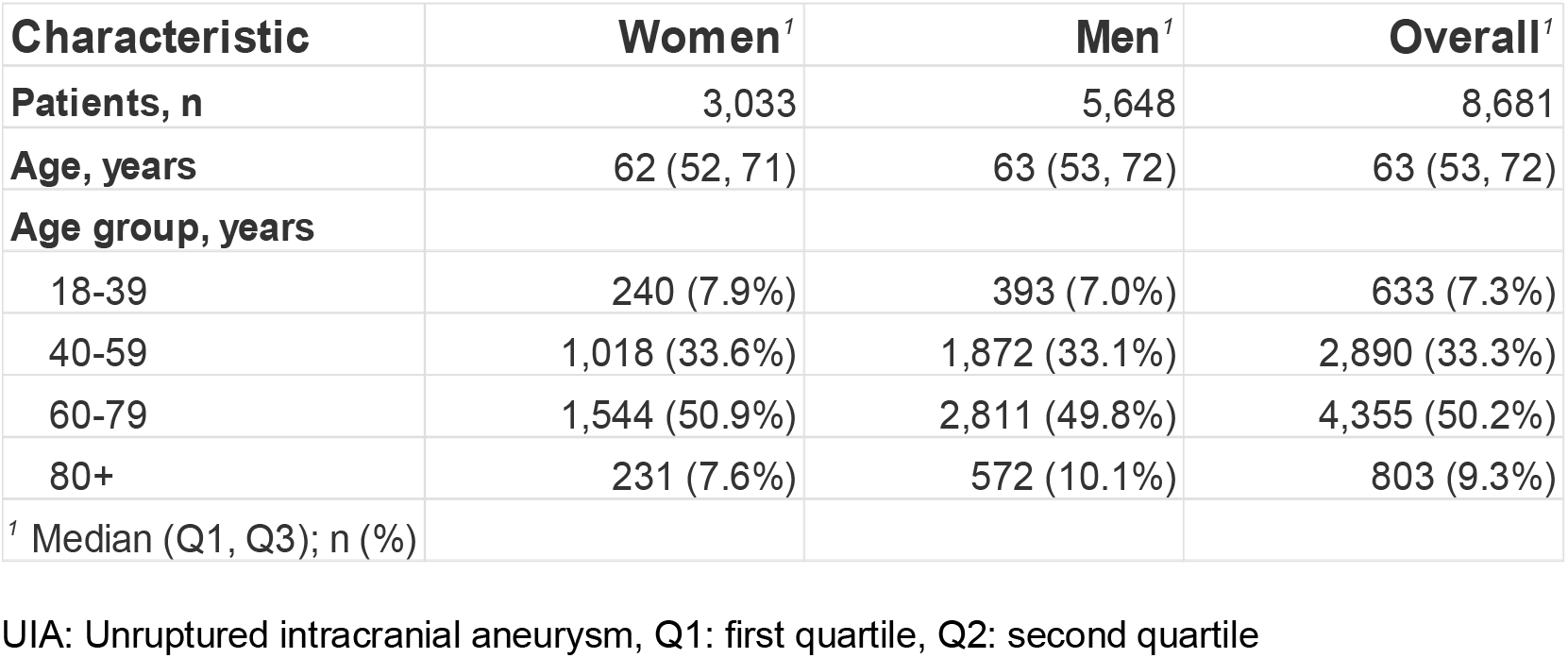
Characteristics of patients diagnosed with UIA, 2000–2020

**Figure 1.**
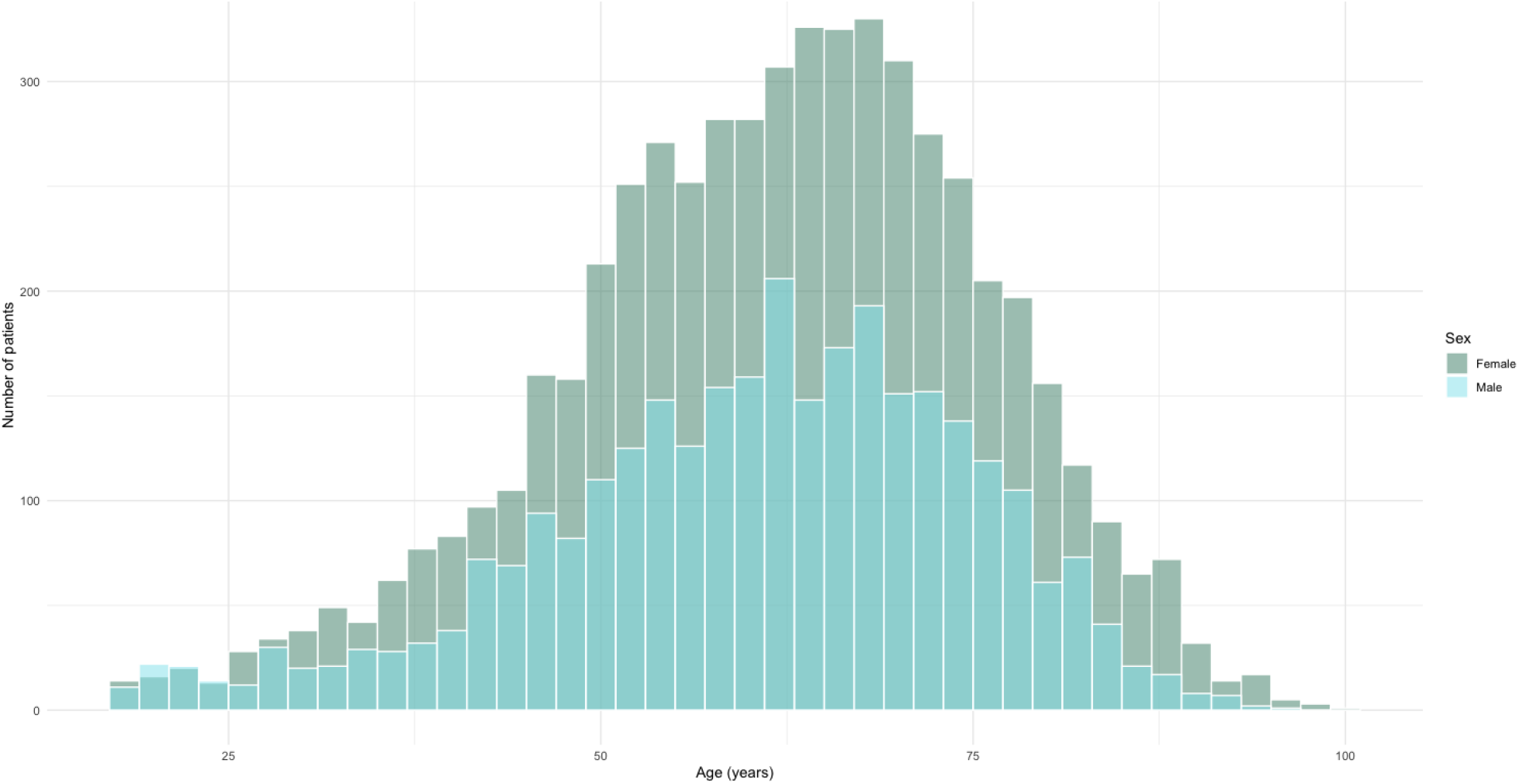
Age distribution of incident cases of UIA by gender in the total sample

Age-standardized incidence of UIA in the total sample rose from 4.6 per 100,000 (95% CI: 3.84– 4.86) in 2000 to a maximum of 7.99 per 100,000 (95% CI: 7.42–8.67) in 2019 (Figure 2) corresponding to a relative increase of 73.7 %. The increase was most pronounced among individuals aged ≥60 years, particularly women, whereas the incidence remained relatively stable among those aged 18–39 years (Figure 3).

**Figure 2.**
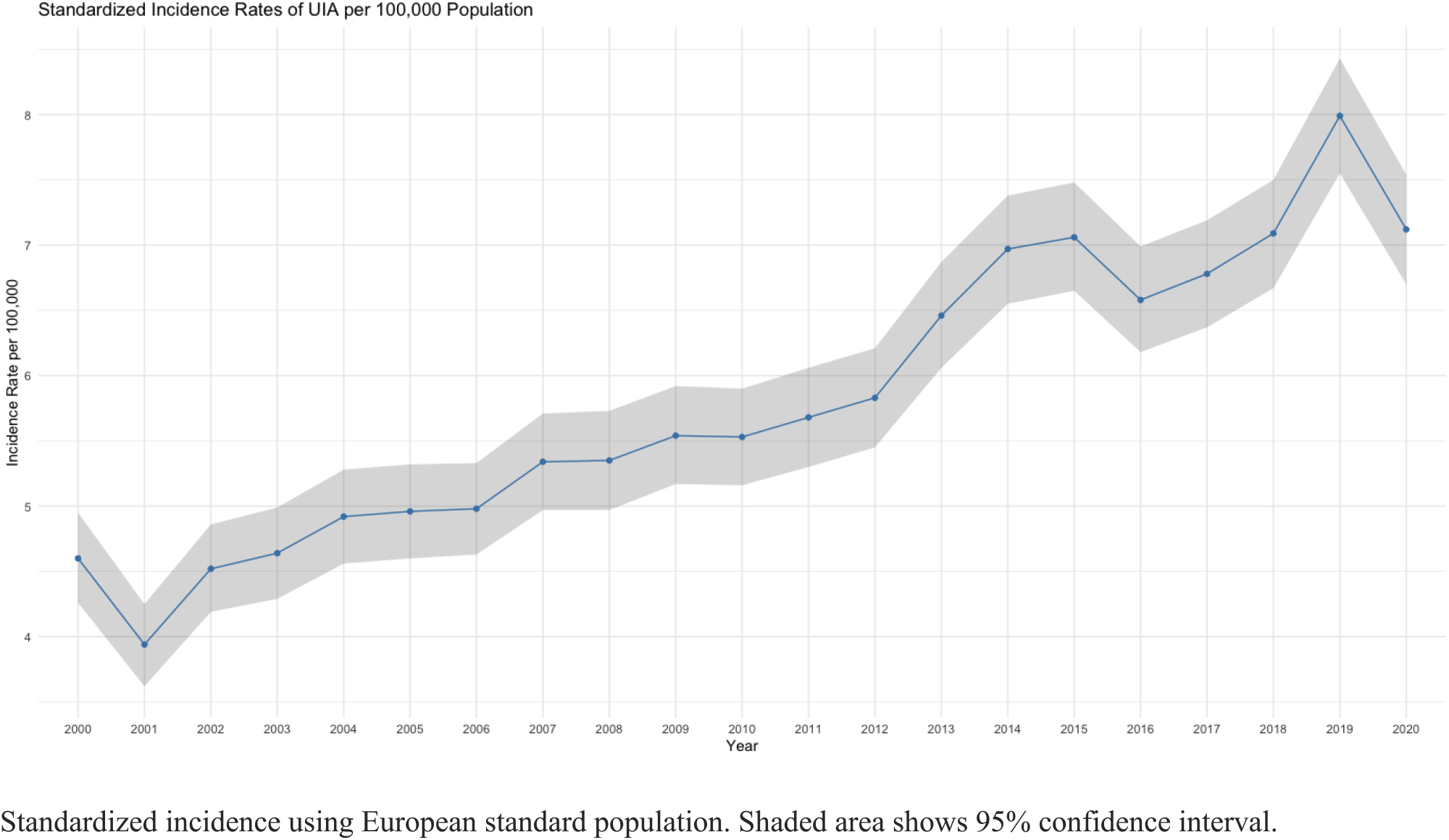
Standardized incidence of UIAs in Sweden from 2000-2020

**Figure 3.**
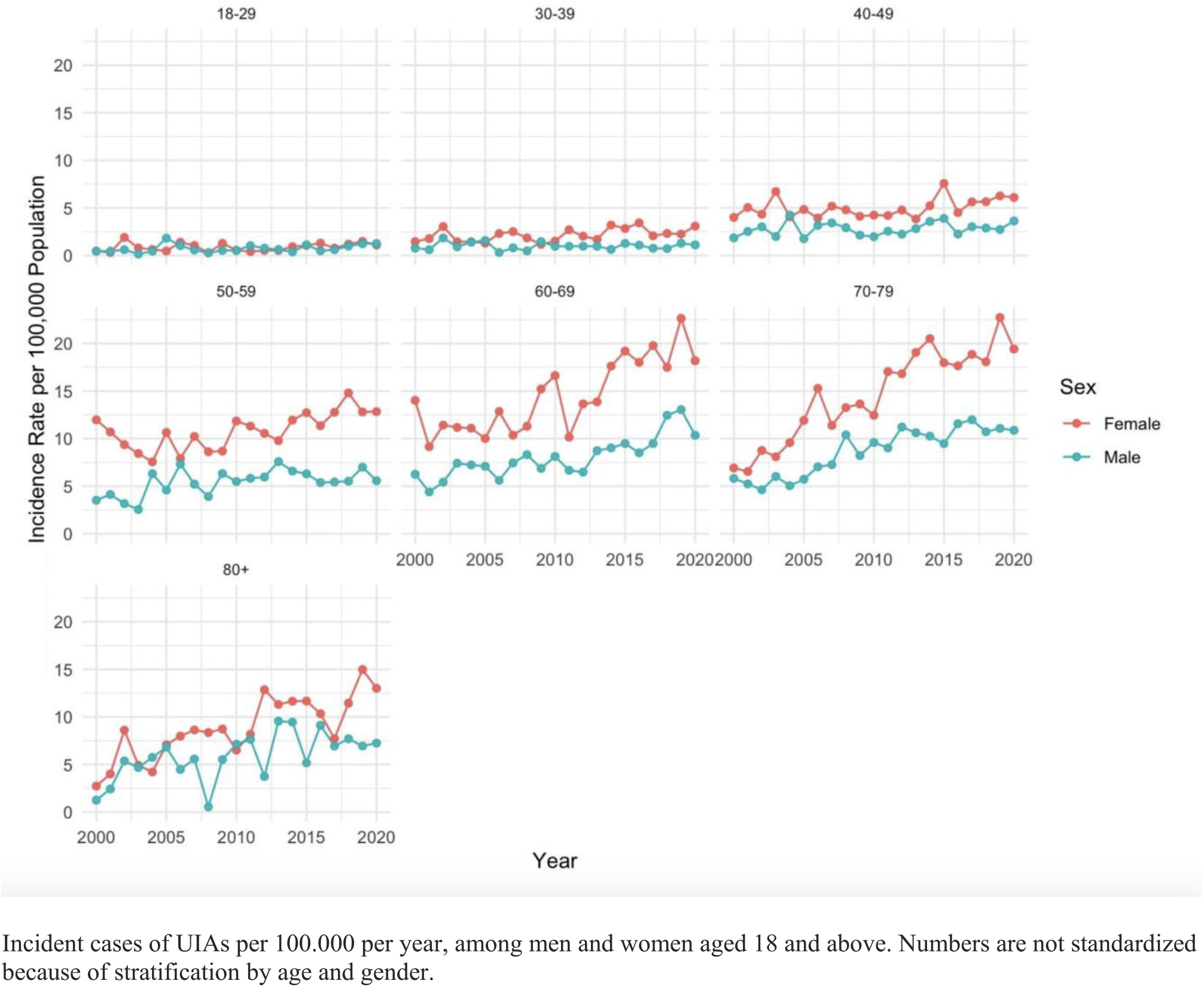
Age-and gender specific incidence of UIA in Sweden (2000-2020)

In contrast, the incidence of SAH in the study sample was more stable during the study period, showing a slight decrease (Figure 4).

**Figure 4.**
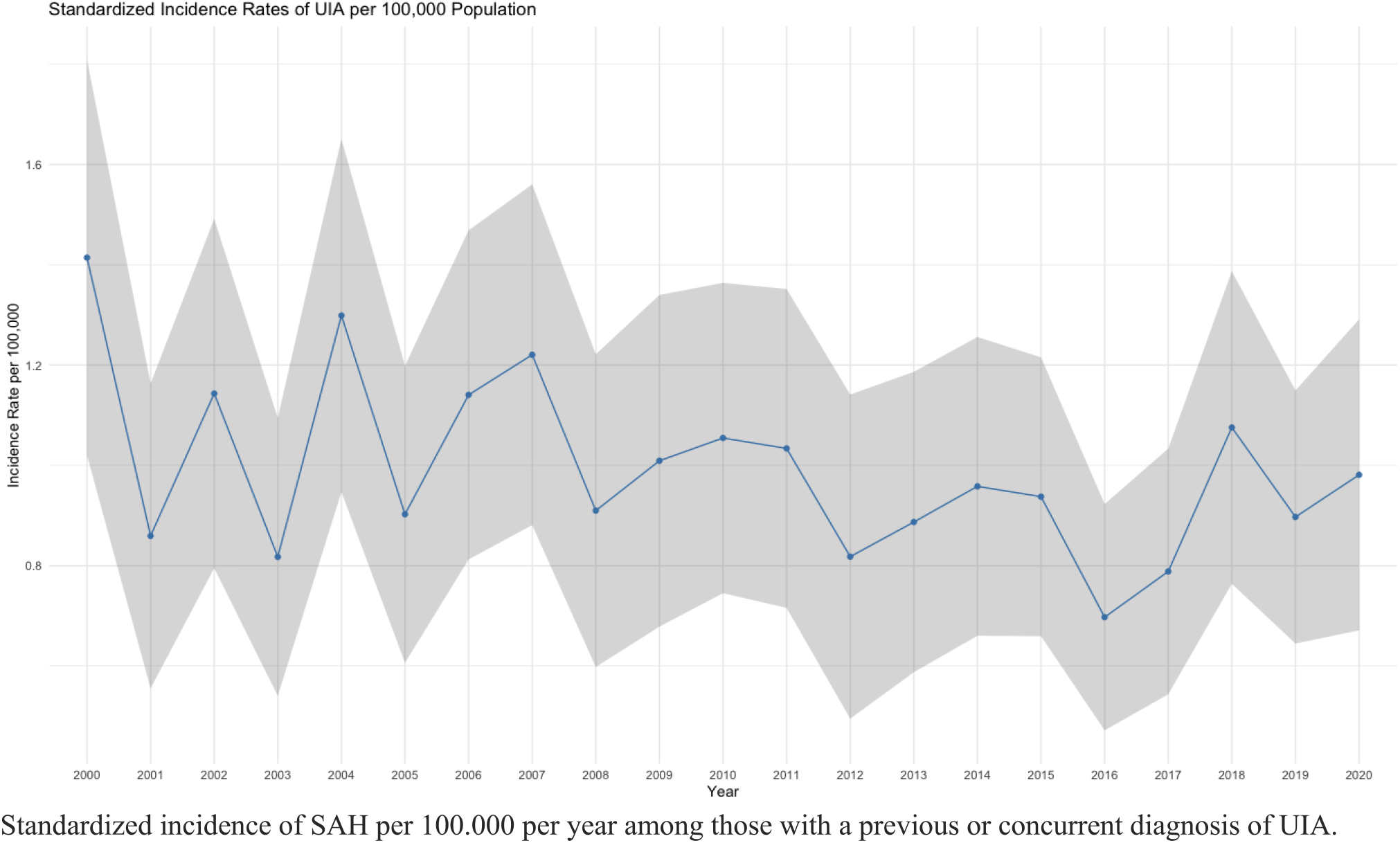
Standardized incidence of SAH in Sweden (2000-2020).

## DISCUSSION

In this nationwide study covering the entire adult Swedish population over a 20-year period we found a substantial increase in the incidence of hospital-based diagnoses of UIAs. The age-standardized incidence, in the total sample, increased by more than 70% during the study, and the most pronounced increase was seen in women and individuals aged 60 years and older. In contrast, the incidence of SAH in the study population was stable over much of the study period - with a slight decrease toward the latter part. These findings together indicate that the number of low-risk aneurysms are increasing considerably, placing a growing burden on health care systems.

### Results in context

In regard to the epidemiology of intracranial aneurysms, much of the past research has focused on the outcome i.e. SAH and there is therefore less data on UIAs. Previous epidemiological studies have estimated the prevalence of UIAs to be approximately 3% in the general population, with higher prevalence among women and older individuals [5]. However, most prior studies have been based on selected cohorts, screening studies, or autopsy series, and there is a relative scarcity of nationwide longitudinal data. Moreover, many earlier investigations employed cross-sectional designs, limiting the ability to assess incidence and temporal trends.

While the incidence of SAH has clearly decreased in several countries, there is less data on how the the incidence of UIAs has changed relative to this [6, 17, 18]. Several previous epidemiological studies have reported increasing incidence rates of UIAs over time, broadly consistent with our findings [19-22]. In contrast, however, several other studies have described stable or even decreasing incidence rates over time [17, 18, 23].

Importantly, results from previous studies have been heterogenous with considerably higher incidence rates compared to this study. For example, we found that the standardized incidence of UIAs was between 4.6 and 7.99/100 000 per year during the study period which is substantially lower than several other studies, from Europe and North America, which have reported incidence rates of approximately 9 to 17/100 000 per year [22-24]. In Korea the incidence has been reported to be as high as 71.4/100 000 per year which is almost 10-fold higher than what we found at the end of our study period [20].

Although the exact mechanisms influencing temporal trends in UIA incidence are unknown, it is possible that a multitude of factors contribute such as local diagnostic practices, healthcare structures, variations in the reporting of diagnoses for quality registries, changes in demographics and risk factor prevalences. Nevertheless, the substantial geographic variability in reported UIA incidence highlights the importance of updated population-based data from different healthcare settings and countries.

Regarding the increase in UIA incidence, the widespread adoption of non-invasive vascular imaging (CT- and MR angiography; CTA, MRA), which have increased drastically, should also be noted. For example, the number of CTAs has increased approximately 10-fold - with a corresponding increase in UIA detection rate of 33 % [25-27]. Taking these data into consideration, it is plausible that increase in incidence of UIAs in this study can largely be explained by increased detection.

The predominance of women and older persons in this sample is in line with well-known epidemiological patterns: Hormonal factors, vascular aging, and sex-specific differences in risk factor exposure have all been proposed as contributing mechanisms [6]. The increasing incidence among older women observed in this study may also reflect a lower threshold for imaging in this group, as well as improved survival from other cardiovascular conditions, leading to a larger population at risk of being diagnosed.

Importantly, the observed increase in UIA incidence occurred in parallel with a slight decline in SAH incidence. Although the decrease in SAH did not reach statistical significance, the divergent trends are important to note. If the increasing UIA incidence represented a true increase in aneurysm formation or growth, a corresponding rise in SAH incidence might be expected. Instead, our findings support the hypothesis that enhanced detection of mostly asymptomatic and potentially low-risk aneurysms has increased the incidence of UIAs without a proportional increase in rupture events. However, it might also be that reduced cardiovascular risk factor burden in the general population together with improved medical management has led to an overall decrease in rupture risk over time, which is consistent with data from most countries [2, 3, 28, 29]. Another possibility is that our findings are due to a combination of the listed factors above.

### Clinical and public health implications

The rising incidence of diagnosed UIAs has important implications for healthcare systems. Increased detection inevitably leads to a larger population of individuals living with a known aneurysm. This places growing demands on surveillance imaging, specialist consultations, and patient counseling. Accurate and updated risk stratification is therefore increasingly critical, particularly in older patients and women, who constitute the majority of newly diagnosed cases. From a public health perspective, our findings underscore the importance of distinguishing between true disease burden and diagnostic activity when interpreting epidemiological trends. While advances in imaging have clear benefits, they also necessitate careful evaluation of downstream consequences, including psychological impact on patients and resource utilization.

### Strengths and limitations

The major strength of this study is the use of nationwide register data with virtually complete coverage of inpatient and outpatient care in Sweden spanning two decades. The universal healthcare system and the high validity of the National Patient Register enhance the generalizability and reliability of the findings. The long study period also allows for robust assessment of temporal trends and age- and sex-specific patterns.

Several limitations should however also be acknowledged. First, registry-based data lack detailed clinical information, including type, size, location and morphology of the aneurysms, all of which are central to rupture risk assessment. Also, we were unable to identify the number of aneurysms in each individual and de novo aneurysm formation in persons with a prior UIA diagnosis. For example, a person with a treated aneurysm who later developed another aneurysm would only have been counted once. Furthermore, coding errors are well known to occur and although the NPR has high diagnostic validity, misclassification is possible, particularly in cases where aneurysmal SAH was not coded together with an intracranial aneurysm diagnosis. It is therefore important to underscore that this study data does not reflect the true SAH incidence in Sweden as we only counted those with a concurrent aneurysm diagnosis. It is likely that many with aneurysmal SAH are only coded only for SAH and not the aneurysm itself. Because of this the actual incidence of SAH in Sweden is probably much higher than reported in this study. On the other hand, including patients with SAH without concurrent aneurysm diagnosis would probably have overestimated the incidence of SAH because of misclassification of non-aneurysmal SAH in the diagnostic coding.

Therefore, including patients with SAH and a concurrent UIA diagnosis likely yields conservative, but more specific estimates. Finally, because most UIAs are asymptomatic, hospital-based diagnoses inevitably underestimate the true occurrence in the population, and the reported incidence should be interpreted as a measure of healthcare-detected disease rather than true biological incidence. Although certain screening studies are more precise and better at assessing the true prevalence of UIAs, they often cannot examine incidence because of the cross-sectional design. Also, hospital-based frequency estimates have the advantage of providing better real-world evidence of the actual utilization of health care resources.

### Conclusions

The incidence of diagnosed unruptured intracranial aneurysms in Sweden increased substantially between 2000 and 2020, particularly among women and older individuals, while the incidence of SAH among those with a diagnosed UIA remained stable or declined slightly. These findings might be more consistent with improved detection and demographic changes rather than a true increase in aneurysm-related disease burden. Future studies integrating detailed imaging characteristics, risk factor data, and long-term outcomes are needed to better inform risk stratification and guide clinical decision-making in the growing population of individuals diagnosed with UIAs.

## Data Availability

All data produced in the present study are available upon reasonable request to the authors.

